# Consistent Pattern of Epidemic Slowing Across Many Geographies Led to Longer, Flatter Initial Waves of the COVID-19 Pandemic

**DOI:** 10.1101/2022.03.31.22273267

**Authors:** Michal Ben-Nun, Pete Riley, James Turtle, Steven Riley

## Abstract

To define appropriate planning scenarios for future pandemics of respiratory pathogens, it is important to understand the initial transmission dynamics of COVID-19 during 2020. Here, we fit an age-stratified compartmental model with a flexible underlying transmission term to daily COVID-19 death data from states in the contiguous U.S. and to national and sub-national data from around the world. The daily death data of the first months of the COVID-19 pandemic was categorized into one of four main types: “spring single-peak profile”, “summer single-peak profile”, “spring/summer two-peak profile” and “broad with shoulder profile”. We estimated a reproduction number *R* as a function of calendar time *t*_*c*_ and as a function of time since the first death reported in that population (local pandemic time, *t*_*p*_). Contrary to the multiple categories and range of magnitudes in death incidence profiles, the *R*(*t*_*p*_) profiles were much more homogeneous. We find that in both the contiguous U.S. and globally, the initial value of both *R*(*t*_*c*_) and *R*(*t*_*p*_) was substantial: at or above two. However, during the early months, pandemic time *R*(*t*_*p*_) decreased exponentially to a value that hovered around one. This decrease was accompanied by a reduction in the variance of *R*(*t*_*p*_). For calendar time *R*(*t*_*c*_), the decrease in magnitude was slower and non-exponential, with a smaller reduction in variance. Intriguingly, similar trends of exponential decrease and reduced variance were not observed in raw death data. Our findings suggest that the combination of specific government responses and spontaneous changes in behaviour ensured that transmissibility dropped, rather than remaining constant, during the initial phases of a pandemic. Future pandemic planning scenarios should be based on models that assume similar decreases in transmissibility, which lead to longer epidemics with lower peaks when compared with models based on constant transmissibility.

**Author summary:** In planning for a future novel respiratory pandemic, or the next variant of SARS-Cov-2, it is important to characterize and understand the observed epidemic patterns during the first months of the COVID-19 outbreak. Here, we describe COVID-19 epidemic patterns observed in the U.S. and globally in terms of patterns of the basic reproduction number, *R*(*t*), using an age-stratified compartmental model. We find that daily death data of the first months of the COVID-19 pandemic can be classified into one of four types: “spring single-peak profile”, “summer single-peak profile”, “spring/summer two-peak profile” and “broad with shoulder profile”. Using the concept of local pandemic time, *t*_*p*_, we show a consistent pattern on four continents of an initial large magnitude and variance in reproductive number *R*(*t*_*p*_) that decreases monotonically and hovers around one for many days, regardless of specific intervention measures imposed by local authorities and without an accompanying decrease in daily death prevalence. We attribute this to significant behavior changes in populations in response to the perceived risk of COVID-19.

## Introduction

The roll out of effective vaccines [1, 2] and the emergence of more transmissible [3–5] and antigenically distinct [6] lineages of SARS-Cov-2 virus [7] marked the end of the global first wave of the COVID-19 pandemic. Over the first year, the COVID-19 pandemic has negatively impacted the health and well being of almost every population around the world. In the absence of an effective vaccine, most countries implemented non-pharmaceutical interventions (NPIs), including travel restrictions, school and work closures, social distancing, contact tracing, quarantining and mask requirements [8–10]. However, the degree of compliance of the population with these measures, and their effectiveness, varied greatly from one setting to another and, even now, is not fully understood [11–15].

The transmission of the SARS-Cov-2 virus is often quantified using the time-varying reproduction number, *R*(*t*), which represents the mean number of secondary cases that a single index case will infect. Many studies have focused on estimating the impact of different interventions on *R*(*t*) under the implicit assumption that interventions are similar between different populations [16–27]. However, the analytical approach in these studies conditions on the assumption that the interventions as measured are the main drivers of changes in *R*(*t*), with transmissibility assumed to be constant otherwise.

In this study, we develop an age-stratified compartmental model [26] with a smoothly varying reproduction number and use it to study the epidemic, from January to October of 2020, in 49 jurisdictions in the contiguous U.S. and 89 locations globally. Our model allows for multiple values of *R*(*t*) and is an extension of our previous work which used a smoothly varying two-value functional form [28–30]. We consider the idea that, whereas epidemic patterns vary from one location to another, trends in *R*(*t*) are consistent if measured relative to “pandemic time”, defined as the time elapsed since the first reported death in a location. Trends in pandemic time *R*(*t*_*p*_) are compared to those of calendar time *R*(*t*_*c*_). Unlike earlier studies, our model fits the inferred daily death and we use a Markov Chain Monte Carlo (MCMC) procedure [31] to fit *R*(*t*) to an increasing number of pandemic days. For each fitting time-window, we analyze the value of *R*(*t*) for the prior two weeks and discuss the results for the U.S. and globally without attempting to correlate any changes in *R*(*t*_*p*_) with specific NPIs.

## Methods

### Data Selection

Many data streams can be used as a measure for the spread of the COVID-19 pandemic [11–13]; however, daily confirmed number of cases, confirmed hospitalizations [1] and cumulative deaths are the most commonly used, and, in particular, these datasets as reported by the Johns Hopkins University (JHU) team [32]. Because of likely biases in the confirmed number of cases (e.g., the large change in test availability over time and the transition to rapid home testing) and the limited availability of hospitalization data, we use the reported confirmed deaths as the most accurate and least biased measure of the pandemic. We note that the death data are also imperfect and may underestimate the true toll of the pandemic (Viglione et al. [33] and Centers for Disease Control and Prevention [34]). We start with the cumulative reported deaths as published by JHU [32] and infer daily deaths for each location. Data for the study were retrieved from the JHU database in November, 2020. Irregularities in reporting may result in negative incidence values for some days and these days are given a weight of zero in our fitting procedure. All other days with non-negative values are given an equal weight of one.

The data were split into two groups: US-states and global. The US dataset included all 48 contiguous states and the District of Columbia. Global data locations were chosen from both country level and administrative level-one divisions (state/province/territory/etc). Sub-country locations include those that appeared in the JHU database on April 31, 2020 (Canada, Australia), but exclude European island territories. Locations in the United States and China were also excluded from the global dataset. After these exclusions, the final list of locations in the global dataset was chosen as the top 110 by cumulative deaths.

Country level population totals and age distributions were taken from the United Nations World Population Prospects 2019 [35]. Male and Female populations are combined and five-year age bins are aggregated to the following 10-year bins: 0-9, 10-19, …, 70-79, 80+. For the United States, state-level populations and age distributions were taken from census data [36, 37], and converted to the same combined-sex and decadal ages format.

### Model and Fits

We use an age-stratified SE[I]_4_RX compartmental model (where X refers to death and four levels of severity for the infectious compartment are included: asymptomatic, flu like, mild and severe) to fit the inferred daily reported death for each location separately. Given the relatively low level of mobility between states/countries during the first nine months of the pandemic, this is a reasonable approximation. The age-specific parameters of the model are taken from [38], and the age specific contact matrix is directly derived from the work of Walker et al. [39] using the”squire” R package available on github: https://github.com/mrc-ide/squire.

In previous studies on influenza and influenza-like-illness [28–30] we used a smoothly varying two-value functional form to describe *R*(*t*). Here we extend this model to an arbitrary number of values:

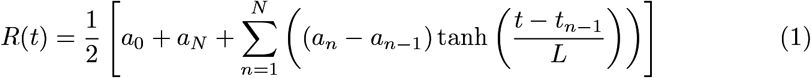

This produces a smooth curve where at roughly time *t*_*n*−1_, the value of *R*(*t*) transitions to *a*_*n*_ with an approximate transition time of *a* ≈ 2*L* days.

For each location, we determine the joint posterior distribution for the model parameters by the fitting the inferred daily reported death using an adaptive step size MCMC [31] procedure with 10^6^ steps. Only the parameters that govern the time variation of *R*(*t*) are optimized (*a*_0_, …, *a*_*N*_ and *t*_1_, …, *t*_*N*−1_) and the timescale of variation is set to approximately seven days (using *L* = 3 in Equation 1). The objective function in the fitting procedure is a Poisson-based Log-Likelihood, and the fitting maximizes the probability that the inferred daily reported death is a Poisson expression of the model daily incidence death. Multiple models of *R*(*t*) (with 2,3, 4 and 5 values, i.e. *N* = 1, …, 4) are fitted to each location and the *AIC*_*c*_ [40] score is calculated for each model. We select the best *N* based on *AIC*_*c*_ score (provided the effective chain size of all the parameters is greater than 50).

We use a simulate-and-recover procedure to validate the model. A known synthetic profile for *R*(*t*) is used to generate synthetic daily death incidence data. The model is fit to synthetic data and the recovered incidence and *R*(*t* profile is compared with the known input (S1 Fig in Supporting Information). The model is able to recover a large variety of synthetic profiles with high accuracy.

To investigate how *R*(*t*) evolved during the course of the pandemic we introduce the concept of “local pandemic time”, defined as the number of days elapsed since the first reported death in a location. For each location, the fitting procedure is repeated using an increasing number of local pandemic days: we start with 30 days of data since the first reported death and increase it to 45, 60, 75, 90 and 105. We also fit and report the reproduction number as a function of calendar time R(*t*_*c*_). Starting with data only until mid-April 2020 and increasing it in five increments of 15 days. This analysis was first applied to the contiguous U.S. (48 states and the District of Columbia) and then extended to 110 locations outside the U.S. The characteristics of calendar and pandemic times *R*(*t*) and the similarities between the U.S. and the world are highlighted in the Results and Discussion sections.

### Data and Code Availability

The dataset used in this study is freely available from JHU [32]. The codes used in this study along with a dataset downloaded on March 25, 2022 and documentation are available from: https://zenodo.org/badge/latestdoi/475441357.

## Results

The U.S. outbreak was first detected in the state of Washington in late February [41]. The next six months of the pandemic can be thought of as a sequence of four state-level archetypal epidemic curves of reported deaths (Fig 1). The first appears as a “spring single-peak profile”. This north-east wave spreads from New York and New Jersey to neighboring states (e.g., Connecticut and Massachusetts), and to the entire north-east corridor (e.g., Rhode Island, New Hampshire, Virginia, the District of Columbia, Maryland, Delaware and Pennsylvania). Overall, during this period, most states in HHS regions 1-3 exhibit the north-east profile with a large peak in daily deaths. We describe the second typical shape we observe as a “summer single-peak profile”. It is observed in a few states that avoided a spring peak but saw their first peak in the summer (e.g. South Carolina, Tennessee, Florida, Texas, Arizona, Arkansas and Idaho). The third typical shape exhibits two peaks, in both the spring and the summer. For example Georgia, Louisiana and Nevada. The fourth typical profile, “broad with shoulder profile”, comes from states that do not exhibit a clear sharp peak (e.g North Carolina, Kentucky, Oregon, Utah, and California) but rather a broad long shoulder-like profile with increases in deaths appearing only in the summer/early fall. Finally, we note that sparsely populated states (such as North and South Dakota), which largely avoided the spring and summer peaks, did not start to show increases in deaths until September.

**Fig 1.**
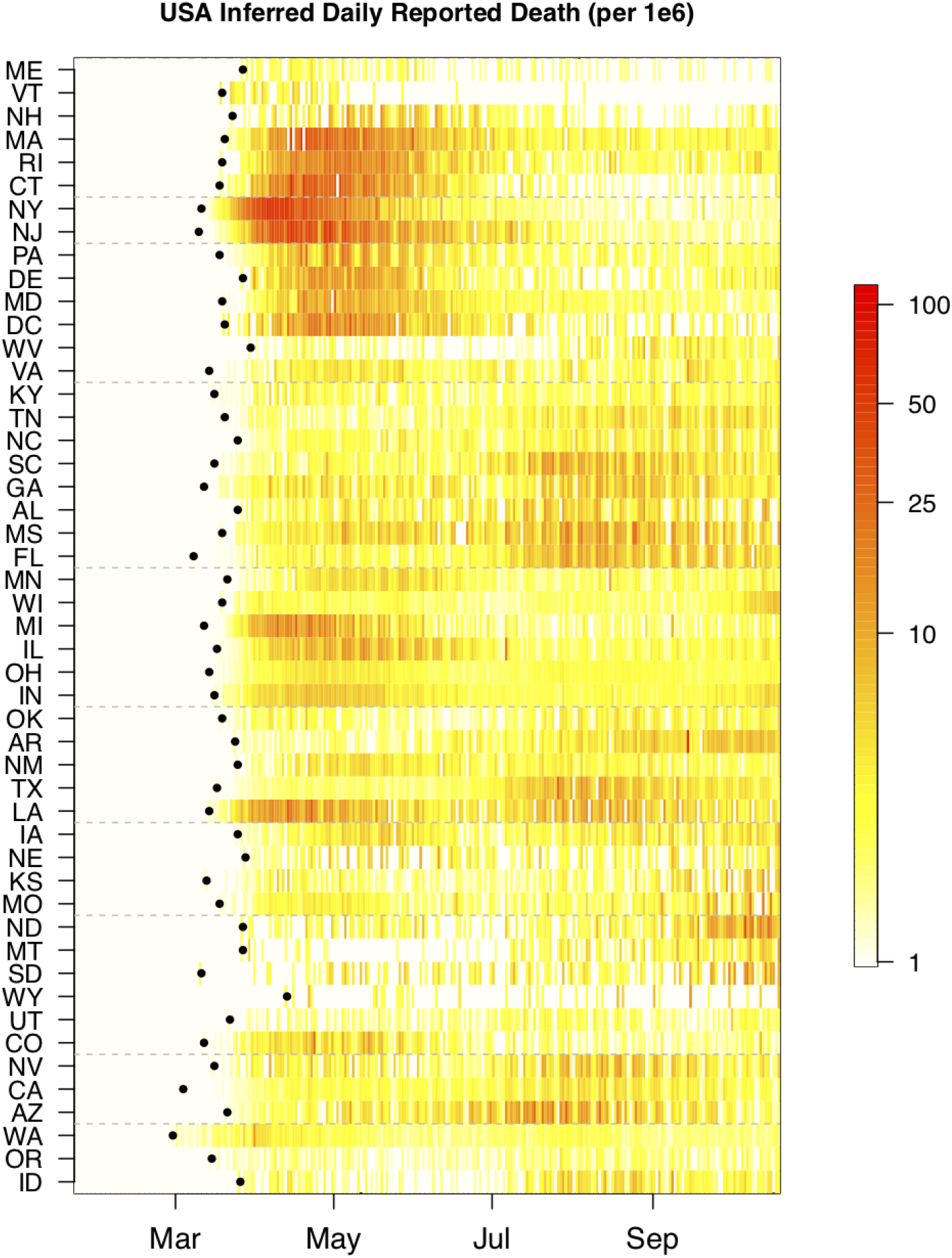
Inferred daily reported deaths (plus one) for the contiguous U.S. From top to bottom, the 48 states and the District of Columbia are ordered by the Human and Health Services (HHS) regions and within each region they are ordered by decreasing latitude. Regions are separated by a dashed grey line. For each location, the date of first reported death is marked with a black dot. Only the first nine months of the pandemic are shown. For clarity, the data are plotted on a log scale and normalized per 10^6^ people.

Individual model results appear in Fig 2 for four U.S. jurisdictions and in S4 Fig of Supporting Information for 15 global locations. The four selected U.S. locations represent the different data profiles described above (summer peak, extended shoulder with summer peak, spring peak, and spring and summer peaks). Similarly, the global locations were chosen to be representative of four continents and all data profiles. The different epidemic profiles lead to different patterns in the time-varying transmissibility when viewed in calendar time. For example, the profile for Pennsylvania starts at *R* ≈ 3 before dropping to just below 1 in May. It maintained that value up to July before rising to ≈ 1.27. In contrast, Texas shows an initially high value (*>* 2.5), then drops rapidly to values just above 1, until late June, at which point it increases to slightly more than 1.5 before returning to values hovering just above 1 for the remainder of the interval. The difference in complexity between the two inferred epidemic profiles is evident in the version of the model with the most parsimonious number of changepoints (see Methods). The number of changepoints with the lowest *AIC*_*C*_ for Pennsylvania was three whereas for Texas it was four. For California and Georgia the number of changepoints was also four. Whereas the details of the patterns in the time-varying transmissibility may vary between locations, we do find that for all four U.S. jurisdictions and most of the 15 locations (exhibiting very different dynamics) the initial value of *R*(*t*) is high (above 3 in Italy, Iran and Sweden) and it decreases to either below or around 1 (dashed grey line). In all cases the model produces a smooth result for *R*(*t*), and with its’ flexible form we are able to fit the variety of profiles described before: early spring only peak (e.g Italy and Sweden), summer only peak (Egypt), spring and summer double peak (Panama and Japan), spring and/or summer and a September/October resurgence (Iran, Russia, Portugal and Armenia), and a broad peak (Bangladesh and Nigeria). The quality of the fits shown in Fig 2 and S4 Fig is representative of the results we obtain for all locations included in this study.

**Fig 2.**
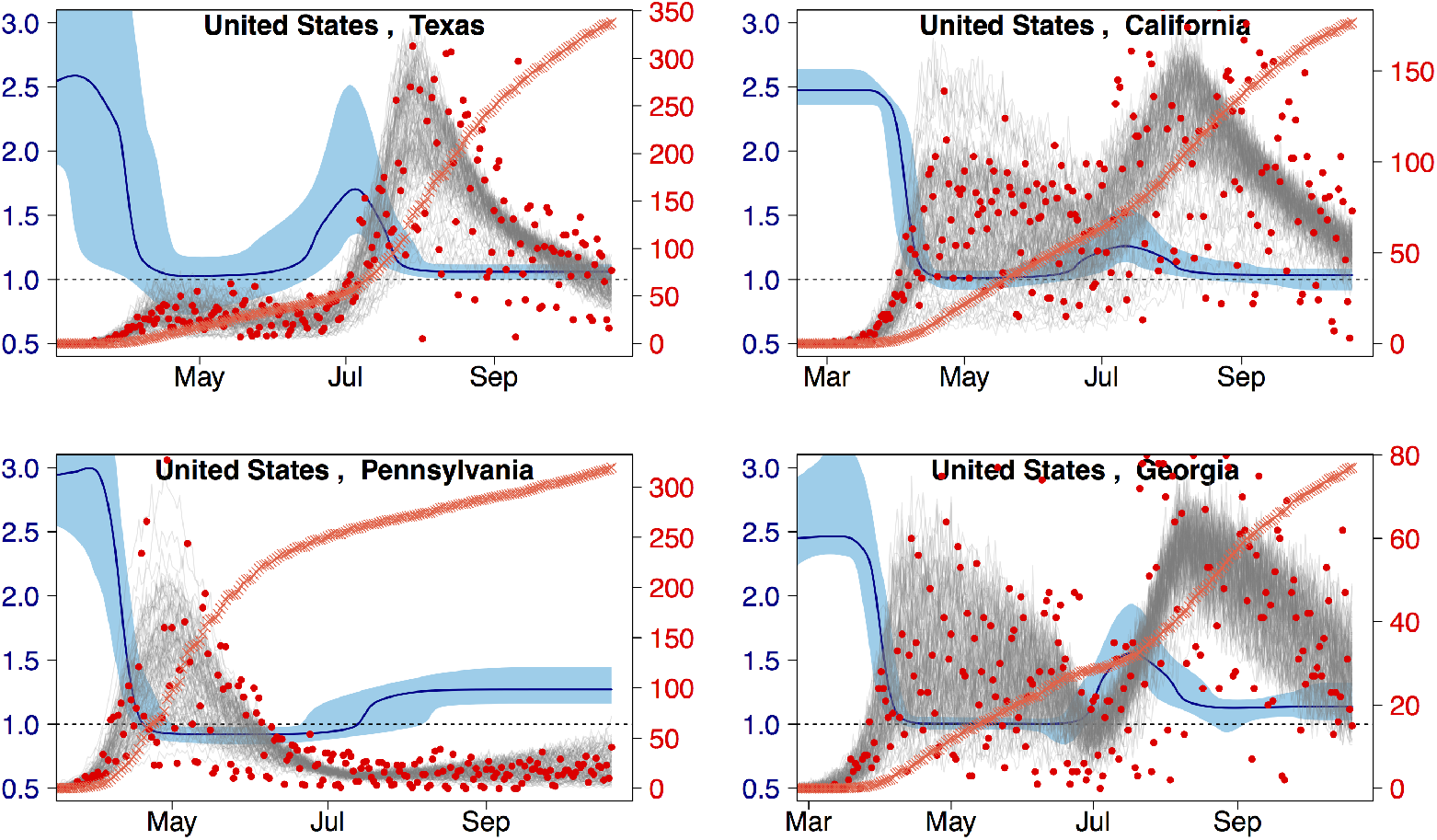
Fitting inferred daily reported deaths. Sample fits to inferred daily reported deaths (plus one) for four U.S jurisdictions red circles and right y-axis. The grey traces are 100 samples from the posterior distribution of the fit and the orange crosses denote the reported per capita cumulative deaths (no y-axis). The median and 95% confidence interval for *R*(*t*) is shown in dark and light blue with the left y-axis. Locations are ordered by decreasing cumulative deaths (not shown).

The distribution of deaths per capita across states in the continental U.S. was stable for the first half of the study. In contrast, the distribution of *R* numbers declined substantially during the same period (Fig 3). We refitted overlapping subsets of the daily death data for the contiguous U.S. starting with data up to mid-April 2020 and increasing in five increments of 15 days (Fig 3 panel(a)). We found initial values of *R*(*t*_*c*_) that were large (mean/median of 2.46/2.33), started to decrease only in May and approached one in June (S1 Table of Supporting Information). We then defined pandemic time *t*_*p*_ – as an alternative for calendar time – as the number of days elapsed since the first reported death in a given population and estimated *R*(*t*_*p*_) the reproduction number as a function of pandemic time (a time shift for each population, Fig 3 panel (b)). Although patterns of *R* in calendar and pandemic time were similar, it dropped more quickly in pandemic time and had a lower variance (S2 Table of SI). The difference between *R* in pandemic time and calendar time was also apparent when we fitted an exponential decay model to both sets of estimates: the model was a much better explanation for the temporal pattern of *R* in pandemic time than it was for calendar time (Tables S3 Table and S4 Table of SI). The daily number of deaths per capita remained far more stable when viewed in both calendar and pandemic time (Fig 3 upper panels). A histogram and heat map representation of the pandemic time evolution of *R* for the contiguous U.S. is presented in S5 Fig of the Supporting Information.

**Fig 3.**
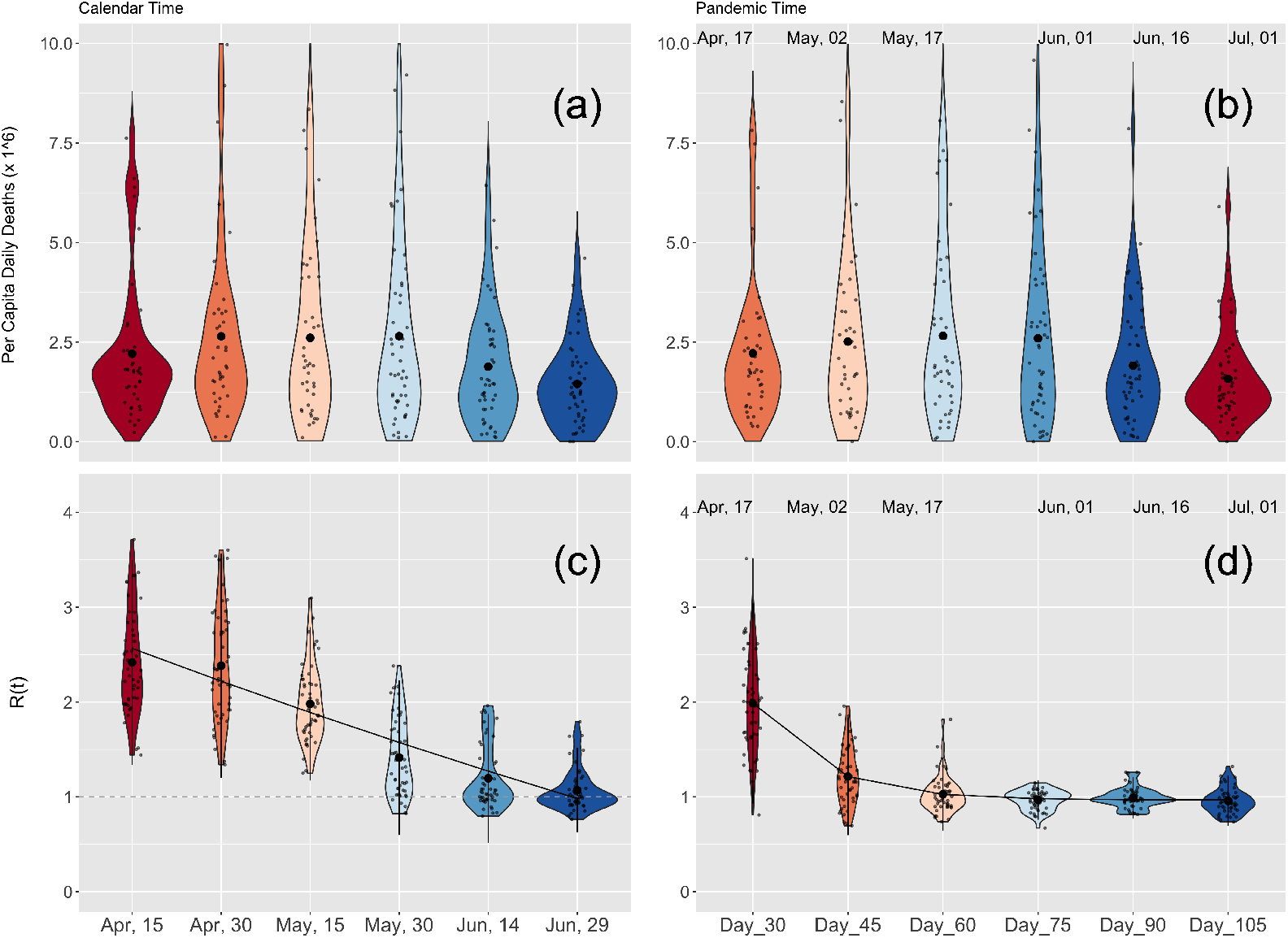
Calendar and pandemic time analysis for the U.S. A violin plot representation of per capita daily deaths for the contiguous U.S. and the distribution of calendar and pandemic times R values for the contiguous U.S. (top and bottom panels respectively). (a) Calendar daily death prevalence calculated using a (centralized) moving average of two weeks using the same dates as in panel (c). (b) Pandemic time daily death prevalence calculated for each jurisdiction using a (centralized) moving average of two weeks around the local pandemic date. (c) Calendar time R(*t*_*c*_) estimated using data from the first reported death in each jurisdiction and up-to the date indicated in the panel. The full black line denotes an exponential fit to the results (see also S3 Table of SI). (d) Pandemic time R(*t*_*p*_) estimated by fitting the first: 30, 45, 60, 75, 90, and 105 days after the first reported death in each jurisdiction. The full black line denotes an exponential fit to the results (see also S4 Table of SI). In panels (b) and (d), the mean date associated with the local pandemic times is indicated above each set of results. In all four panels, to increase readability, a jitter is applied to the displayed data points.

We repeated our analyses for populations outside the U.S. We grouped 110 global locations by continent (Africa, Americas, Asia-Oceania, Europe) and again ordered them by decreasing latitude (S2 Fig of Supporting Information). Here, too, we observed rich dynamics in the timeseries of reported deaths on all continents, with all profile types present that were observed in the contiguous U.S. (see above). In the Americas, the spring single peak profile was observed only in Canada and Ecuador whereas most states in Central and South America showed either the summer or (late summer peak) single peak. The two countries with a large number of deaths in South America (Brazil and Mexico) also showed a large wide peak that extended over more than three months. The data for Asia-Pacific showed an increase in death in most places only in June (e.g., the single summer peak profile). However, Iran was a notable exception, showing the spring-summer double peak profile (with the first peak having already occurred in April) followed by a third resurgence in September. In Australia (and particularly the state of Victoria) we see the single summer peak profile (with the peak in death occurring in August/early September). Examples for the summer single peak profile are Bangladesh and Saudi Arabia, which largely avoided any excess death during the spring. The data for Europe showed clear regional grouping with Italy and Spain leading the spring single peak followed closely by most of the larger European countries.

For both calendar and pandemic time we found similar trends to those observed in the contiguous U.S. (Fig 4 and S5 Table and S6 Table of SI). For consistency, our analysis of *R* for calendar and pandemic time included only 89 out of the 110 locations with two or more weeks of data for all dates after April 15, 2020. For the calendar *R*, the initial values in mid-April were above two and had a large variance (mean/median and standard deviation of 2.48/2.30 and 0.76). As with the U.S. populations, the decline in magnitude and variance of the calendar *R* was slower than the exponential decline of *R* in pandemic time R, which hovered around a value of one for many days. However, the rate of decline in transmissibility in pandemic time was slower than that for the contiguous U.S. (compare the top right panels of 3 and 4, and S2 Table and S6 Table, and S4 Table and S8 Table). We have verified that our conclusions for the calendar and pandemic *R*(*t*) do not change when more global locations are included in the analysis (S3 Fig in Supporting information). For the contiguous U.S., here, too, we found that the daily number of deaths per capita remained far more stable than transmissibility when viewed in both calendar and pandemic times (Fig 4 lower panels). A histogram and heat map representation of the pandemic time evolution of *R* is presented in S6 Fig of the Supporting Information.

**Fig 4.**
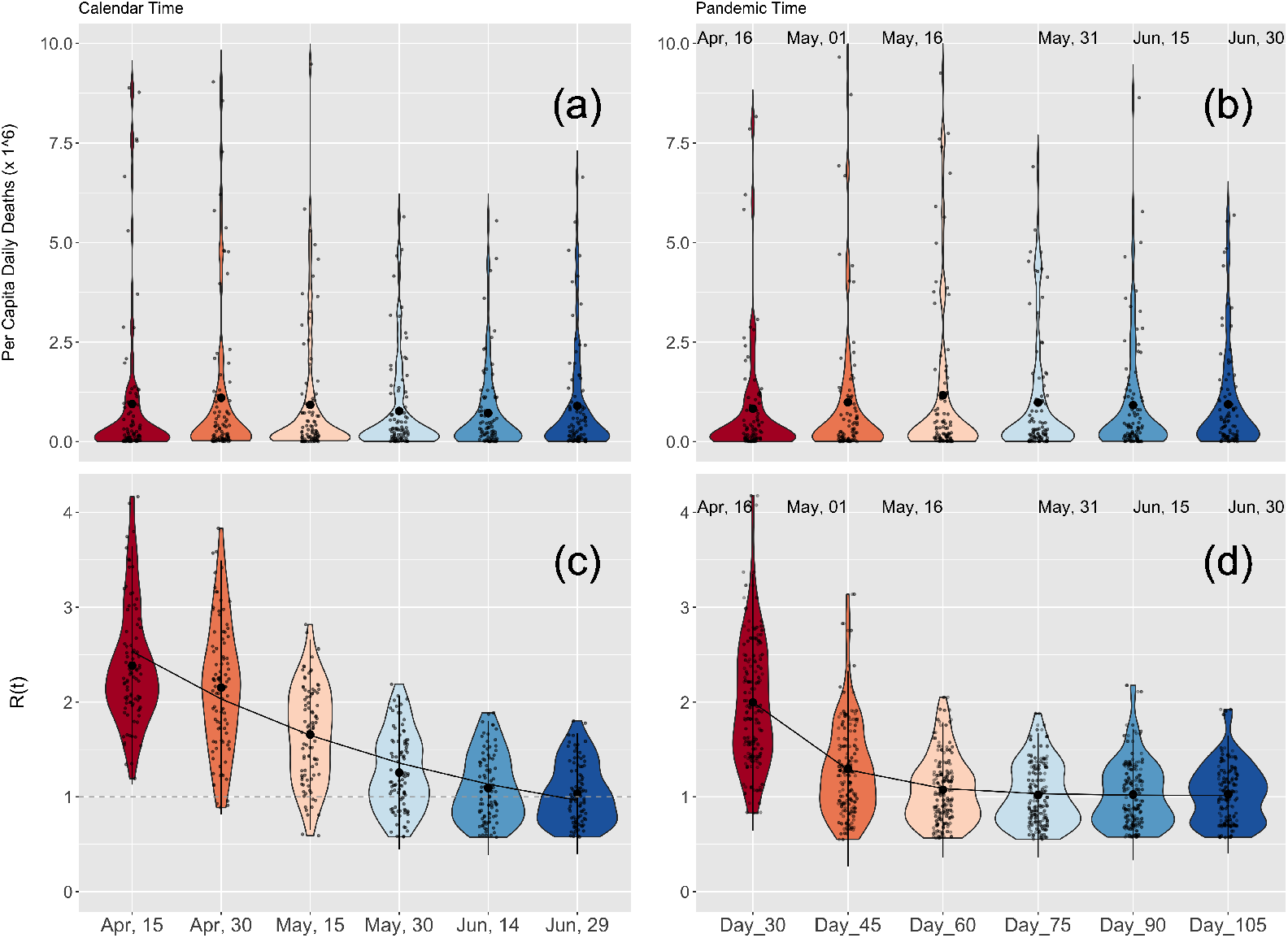
Calendar and pandemic time analysis for the world. Same as Fig 3 but for 89 world locations. (See Tables S7 Table and S8 Table of SI for the exponential fit parameters.

## Discussion

We describe a consistent pattern of an initial high value of *R*(*t*_*p*_) that falls steadily during the first few months after deaths are recorded in a population. Using an age-stratified compartmental model fit to different populations around the world. We found that initial values and variance of *R*(*t*_*p*_) were large. However, as the pandemic progressed, the magnitude and variance of *R*(*t*_*p*_) decreased monotonically, eventually hovering around one for a prolonged period. While the magnitude and variance of *R*(*t*_*p*_) estimated from deaths decreased consistently across the globe, the daily number of deaths themselves did not. In contrast, as a function of calendar time, the initial value of *R*(*t*_*c*_) was larger (a mean value of 2.47 and a variance of 0.37/0.58 for the U.S./world) and the decrease in magnitude was slower, with a lower reduction in the variance.

In contrast with many other studies [15–27, 42–45], we did not explicitly consider individual interventions as explanations for reducing values of *R*. Rather, we investigated overall trends in *R*. This, we believe, may help guide higher-level planning for the next pandemic caused by a severe respiratory pathogen. We found a common pattern of a reduction in transmissibility during the initial period, rather than constant transmissibility, of which there were no obvious examples. Therefore, regardless of the specific intervention plans in place for individual countries, we suggest that initial planning for future similar pandemics should not be based on assumptions of prolonged constant transmissibility driving a rapid peak and the development of population immunity.

Our study relies on a number of potentially important assumptions, approximations and limitations. First, we chose the daily deaths as the dataset to fit to the model, inferring it from cumulative confirmed deaths, as opposed to using other data such as cases, which have been associated with known and considerably larger biases. However, while the deaths are likely a more reliable measure of the pandemic than cases they are far from perfect. Different states and countries use different criteria when registering deaths (for example, some report only confirmed deaths while others report both probable and confirmed deaths) and have different delays in reporting. Additionally, the reported numbers have been shown to be lower than the true toll of the pandemic (see e.g [33, 34, 46–49]).

Our age-stratified compartmental model treats each population separately and ignores travel and importation of cases from other locations. While both global and local travel were significantly disrupted in 2020, they were responsible for the initial spread of COVID-19, and continue to play a role during latter periods. In this work, we ignored the initial introduction of cases to a location and focused on the dynamics of the virus following its introduction to a population. Also, we assumed the same quality of care over time at all locations. In practical terms, in the model, we assume that the probability of death from COVID-19 depended only on age during this period prior to wide spread vaccination. In reality, treatments for severe patients have improved over time, and they varied from one location to another. (We note however, that the first treatment for COVID-19, the antiviral Remdesivir, was fully approved by the FDA only on October 22, 2020 [50], which coincides with the end of the time frame of this study, and likely had little impact in most places.)

We note that a large number of studies have used available databases of interventions [8–10], coupled with statistical methods, to estimate the impact of different interventions on the time-varying reproduction number [15–27, 42–45]. In an extensive study, Liu et al. [16] applied statistical methods to 13 categories of NPIs using confirmed cases data from January to June of 2020 from 130 countries. They concluded that there is a strong association between school closure and internal movement restrictions on the reduction in *R*_eff_(*t*), which was taken from EpiForecast [51] and based on the Cori method [52]. Another major statistical study by Haug et al. [15] used a large database of interventions [8] and multiple statistical methods to quantify 6,068 NPIs in 79 locations on *R*_eff_(*t*), concluding that less intrusive and costly NPIs can be as effective as more intrusive ones. A Bayesian hierarchical model was used by Brauner et al. [21] to study 8 NPIs in 41 countries using national case and death counts (January-May 2020), concluding that the banning of large gatherings and school closures had a large relative reduction on *R*(*t*).

Although the methods used here have some similarities with other published studies, the use of compartmental models with a time-varying reproduction number has been more limited. In the initial phase of the pandemic, Linka et al. [42] used a two-value time-varying reproduction number and an SEIR model to study the correlation between the reproduction number of COVID-19 and public health interventions in Europe. The reproduction number was fit using a machine learning (ML) procedure and the authors found a strong correlation between passenger air travel, driving, walking and transit mobility and the effective reproduction number (with a long time delay of about 17 days). In another study Dickman [43] developed a deterministic SEIR model without age or spatial structure but with a time-dependent reproduction number. The model allowed the transmission parameter, *β*, to have three piecewise constant values and the parameters of the model were obtained by least squares fitting to the reported cumulative confirmed number of cases at a large number (160) of locations, providing insight into the initial decrease in the value of the reproduction number. Anderson et al. [44] studied the effect of social distancing measures using early (March-April 2020) case-count data from British Columbia and five other jurisdictions. They developed an SEIR model with physical distancing compartments that contribute a reduced amount to the force of infection. In this model, the fractional reduction to the force of infection is due to increase in physical distancing and it is allowed to linearly change, over the course of one week, from 1 (no physical distancing) to a final value which they estimate using a Bayesian statistical model. Duque et al. [45] describe an age and risk stratified SEIR-style model with a transmission parameter that is reduced during stay-at-home/work-safe-order time periods and use it as part of an optimization framework that seeks to minimize lockdown days while preventing healthcare surges. Weitz et al. [53] developed an SEIR model with “short-term awareness of risk”, that depends on the death rate, and showed that this feedback mechanism can result in highly asymmetric epidemic curves and death time-series with extended plateau periods. However, this model was not used to fit any data. Implicit in all of these studies is the assumption that interventions are similar between different populations and that they can be correlated to changes in *R*(*t*). The main differences between our work and these studies are that we did not attempt to correlate changes in *R*(*t*) with any specific NPIs and we used an age-stratified compartmental model with a flexible, smoothly varying, time-dependent reproduction number.

Whereas the conclusions of our study do not directly contradict any of these previously published results [15–27, 42–45], our approach of using the dynamics and prevalence of COVID-19 to understand patterns in *R*(*t*) is unique in several ways, and particularly through the introduction of “local pandemic time”. Changes in the time-dependent transmission are driven by many factors that vary from one location to another. For example, some countries (e.g. South Korea [54, 55]) were effective in quickly tracing and isolating new COVID-19 cases and some (e.g. Australia [56] and New Zealand [57]) imposed long lockdowns. Many countries kept K-12 schools closed for the entire time period of this study [58, 59], whereas a few, e.g. Denmark [60], were successful at re-opening their education system without significantly increasing community transmission. Avoiding (or closing) indoor settings and imposing masks requirements varied from country to country (and state to state) [61] as did the enforcement and public adherence to these guidelines/orders [62]. The ability to isolate and protect elderly and otherwise vulnerable populations significantly impacted the cumulative confirmed death in many countries. For example, European countries, including the United Kingdom, failed to protect their senior population [63]. The methodology presented here avoids the need to estimate the impact of any of these factors on the time-varying reproduction number. Yet, at the same time, it also provides a starting point for attempting to understand these impacts in the future.

Our study focuses on the first wave of the COVID-19 pandemic. The monotonic reduction in the time-varying reproduction number persisted in the U.S. and globally for many days but, during the last three to four months of 2020, it began to increase in nearly all locations in the northern hemisphere, as well as many in the southern hemisphere (e.g. South Africa). This increase marked the beginning of a global second wave and was due to climatic effects and the spread of the novel Alpha (B.1.1.7) and Beta (B.1.351) variants first detected in south-east England and South Africa in September 2020 [7], with the former estimated to be greater than 50% more transmissible than pre-existing variants [3–5]. Similarly, the next wave of COVID-19 infections was driven by the novel Delta (B.1.617) variant and the last by Omicron (BA.1 and BA.2) [7, 64, 65]. All three variants, and in particular Omicron, spread faster than the variants dominant during the time frame of this study. To describe the emergence, and rise to dominance of a new variant, compartmental models will need to be extended to include at least two strains with different transmissibility. These subsequent waves resulted in a massive increase in cases, hospitalization and deaths that eclipsed that of the first wave, in spite of a global vaccinations effort and the development of multiple new treatments (e.g. monoclonal antibody treatments).

General properties of the initial wave of the COVID-19 pandemic will likely remain a topic of considerable interest for many years [66]. In future studies, we plan to extend the framework developed here. Should other datasets (e.g., case counts and/or hospitalizations) become less biased and more available, we will incorporate them into the objective function we fit. Our model can also be made more flexible by including differences in quality of healthcare over time and location, which will allow for coupling between geographic regions [29]. Our previous work on forecasting ILI in the U.S. [28–30] highlights the important role that spatial coupling can play in respiratory disease transmission, and which we anticipate will become increasingly more important as travel restrictions have been relaxed and movement between states, countries, and continents significantly increases.

## Data Availability

The codes used in this 105 study along with a dataset downloaded on March 25, 2022 and documentation are 106 available from: https://zenodo.org/badge/latestdoi/475441357.

## Acknowledgments

We thank the CDC and the MIDAS coordination center for organizing and hosting helpful weekly COVID-19 related conference calls. This work was supported by the National Science Foundation Award number 2031536, and by Cooperative Agreement number NU38OT000297 from The Centers for Disease Control and Prevention (CDC) and CSTE and does not necessarily represent the views of CDC and CSTE.

## Supporting information

**S1 Fig.**
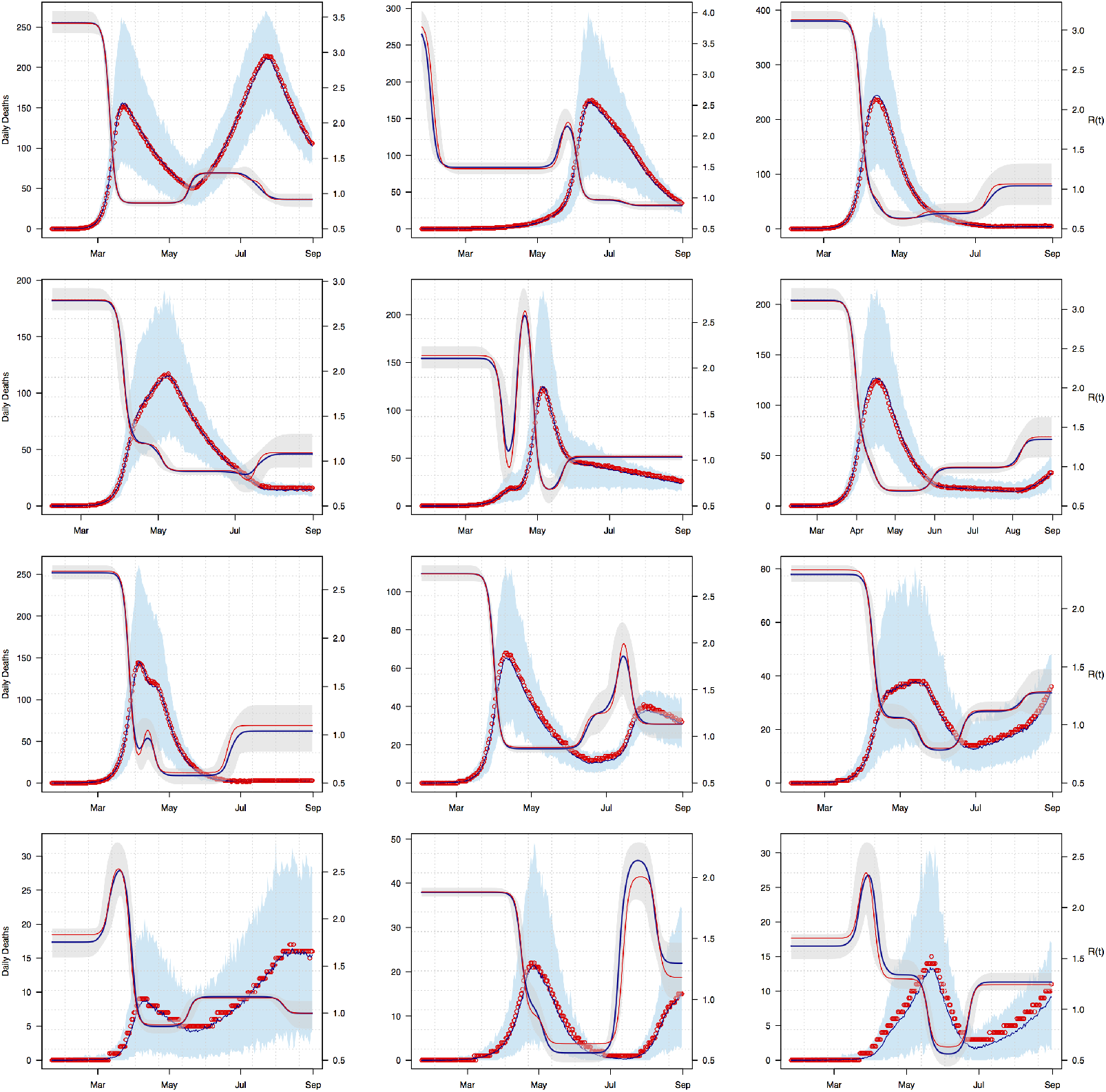
Simulate and Recover. Sample fits to synthetic data generated using known R(t) profiles. The synthetic daily death (red circles and left y-axis) is generated using known synthetic *R*(*t*) profiles (red line and right y-axis). The median result of the fit is shown in blue and the shaded light-blue is the 95% CI. The recovered, median, *R*(*t*) is also shown in blue along with the 95% CI in light-grey.

**S2 Fig.**
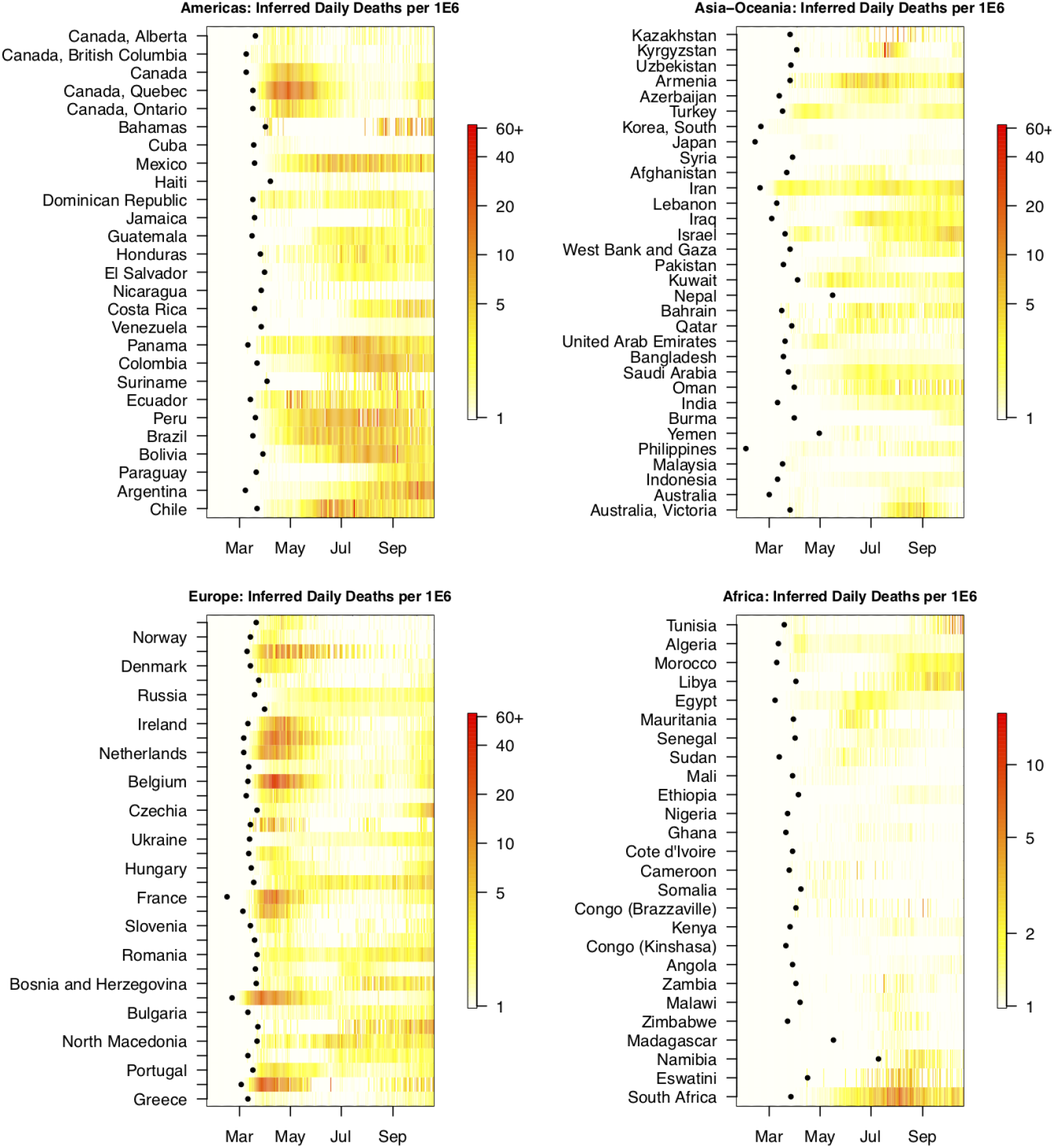
Global Inferred per capita daily death. Inferred per capita daily reported deaths for 120 locations (other than the U.S.) grouped by continent (with Australia grouped with Asia). To balance the number of locations on each continent, here we show more locations than used for the analysis of *R*. (See text for more detail.) Within each panel locations are ordered by decreasing latitude from top to bottom. For each location, the date of the first reported death is marked with a black dot. For clarity, data is shown on a log scale and saturated at 60 for all panels other than the bottom right.

**S3 Fig.**
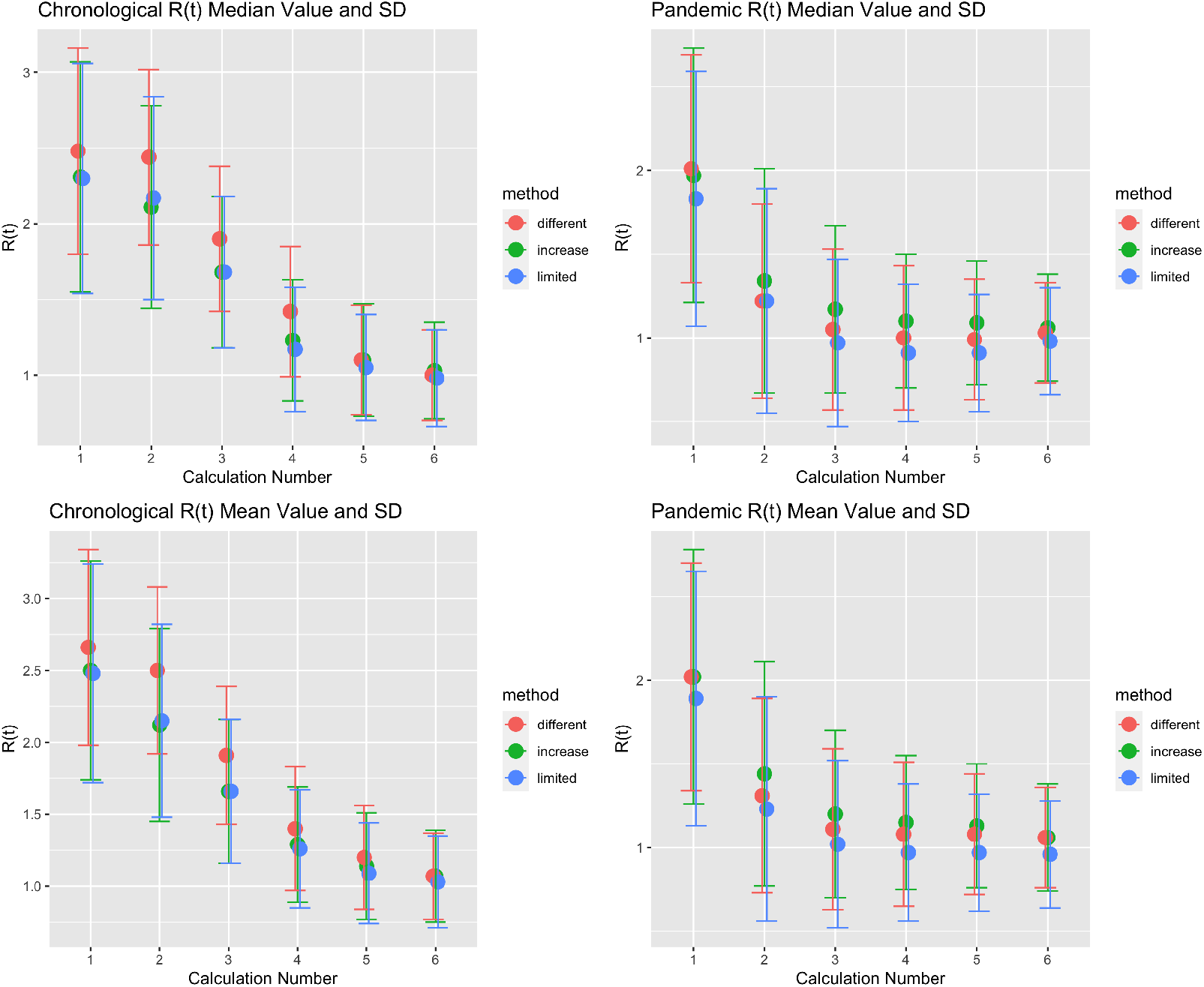
R values sensitivity analysis. Median/mean (top/bottom row) calendar/pandemic (left/right column) R values and their standard deviations inferred using different selection procedures for 89-110 global locations. Limited: for both calendar and pandemic analysis use the same limited subset of 89 locations for which there was two weeks or more of daily inferred death data for the first calendar time calculation. Increase: use the same subset of locations for the six calendar and pandemic (R(*t*_*c*_) and R(*t*_*p*_)) analysis, but allow the number of locations to gradually increase from 89 to 110 as more locations have sufficient calendar data. Different: the calendar analysis includes the subset of 89 locations that had sufficient data at the time of the first calendar calculation and the pandemic analysis includes all 110 locations that have sufficient data for at all six pandemic times.

**S4 Fig.**
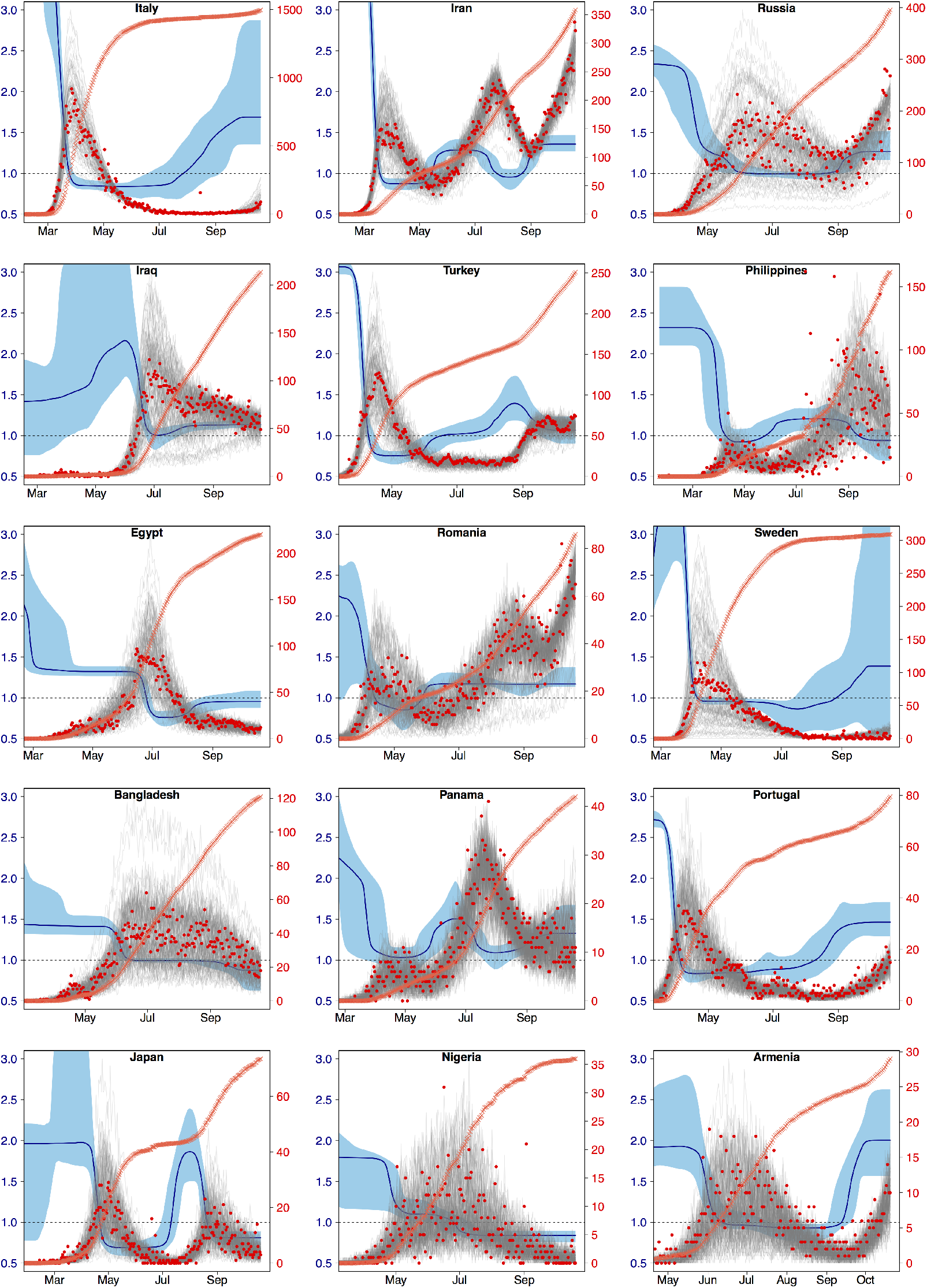
Daily inferred death fits. Sample fits to inferred daily reported deaths (red circles and right y-axis) from 15 countries. The grey traces are 100 samples from the posterior distribution of the fit and the orange crosses denote the reported per capita cumulative deaths (no y-axis). The median and 95% confidence interval for *R*(*t*) is shown in dark and light blue with the left y-axis. Locations are ordered by decreasing cumulative deaths (not shown).

**S5 Fig.**
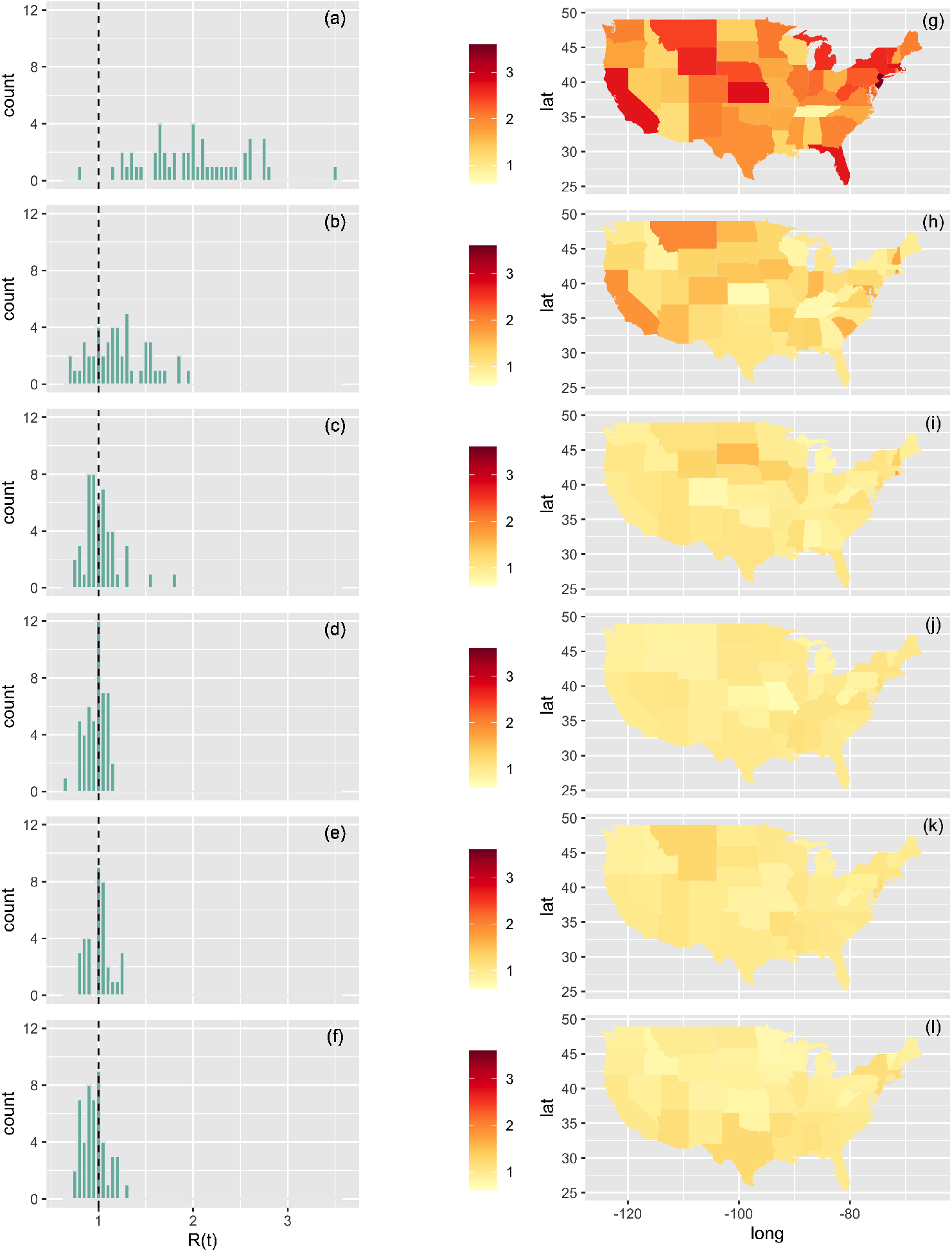
Left column: histogram plots of the pandemic time R(*t*_*p*_) values for the contiguous U.S. as calculated by the model by fitting the inferred daily reported deaths using 30, 45, 60, 75, 90 and 105 days (panels (a) to (f)) since the first reported death in each location. The black vertical dashed line is at *R*(*t*_*p*_) = 1. Right panel: a heat map representation of the data showing the value for each of the 49 contiguous jurisdictions.

**S6 Fig.**
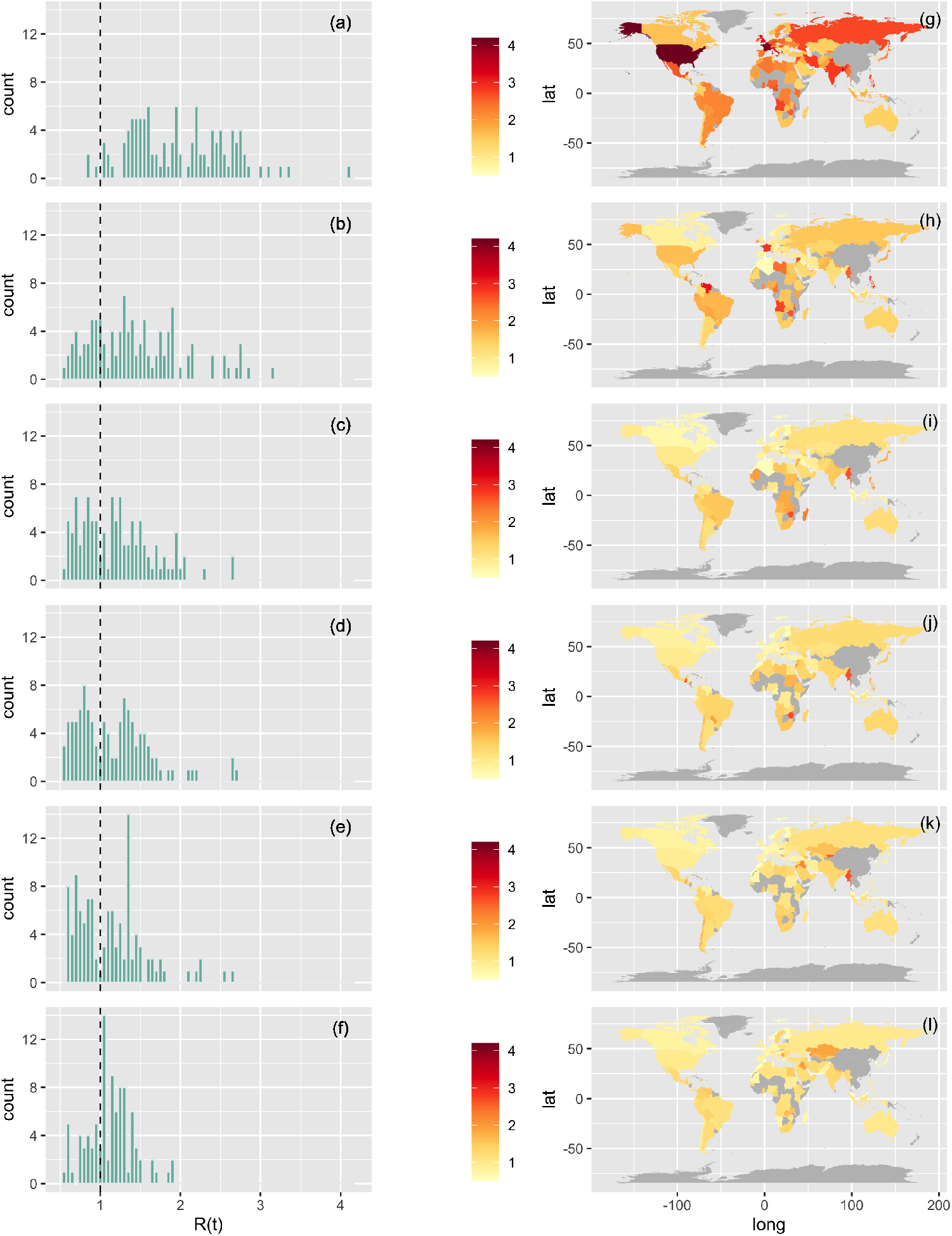
Same as S5 Fig but for 110 world locations. For clarity the entire U.S. is treated as a single country in these maps and we display results for more locations than the 89 discussed in the text and tables.

**S1 Table.** Calendar time reproduction number. Estimated R(*t*_*c*_) values for the contiguous U.S. (49 jurisdiction) as a function of calendar time. Numbers in parentheses denote the 95% confidence interval of the mean and median values and SD denotes standard deviation.

| Date | Mean | Median | Standard Deviation | #Below one |
| --- | --- | --- | --- | --- |
| 04-15-2020 | 2.46(2.28-2.64) | 2.33(2.15-2.54) | 0.61 | 0 |
| 04-30-2020 | 2.38(2.21-2.55) | 2.25(2.13-2.71) | 0.59 | 0 |
| 05-15-2020 | 1.98(1.87-2.10) | 1.91(1.78-2.09) | 0.41 | 0 |
| 05-30-2020 | 1.41(1.30-1.53) | 1.32(1.15-1.58) | 0.41 | 5 |
| 06-14-2020 | 1.20(1.10-1.29) | 1.04(1.00-1.13) | 0.34 | 17 |
| 06-29-2020 | 1.07(1.01-1.14) | 1.00(0.95-1.05) | 0.22 | 23 |

**S2 Table.** Pandemic time reproduction number. Same as S1 Table but as a function of local pandemic time R(*t*_*p*_).

| Days since <sup>st</sup> death | Mean | Median | Standard Deviation | #Below one |
| --- | --- | --- | --- | --- |
| 30 | 1.99(1.84-2.14) | 2.00(1.73-2.11) | 0.53 | 1 |
| 45 | 1.22(1.13-1.30) | 1.19(1.08-1.28) | 0.31 | 13 |
| 60 | 1.03(0.97-1.08) | 1.00(0.94-1.06) | 0.19 | 25 |
| 75 | 0.97(0.94-1.00) | 1.00(0.94-1.02) | 0.10 | 27 |
| 90 | 0.99(0.96-1.02) | 0.97(0.96-1.01) | 0.11 | 30 |
| 105 | 0.96(0.92-1.00) | 0.94(0.90-0.99) | 0.13 | 33 |

**S3 Table.** Exponential Fits to R. Results of exponential fit *R* (*t*) = *R*_∞_ + (*R*_0_ − *R*_∞_) *e*^−*αt*^ to calendar R(*t*_*c*_) values for the 49 U.S. jurisdictions.

| Parameter | Estimate | Standard Error | $Pr(> \vert t\vert )$ |
| --- | --- | --- | --- |
| $R_\infty$ | -5.37 | 10.98 | 0.62 |
| $R_0$ | 2.57 | 0.061 | $< 2^{-16}$ |
| $\alpha$ | 0.003 | $6.6 * 10^{-4}$ | $1.97 * 10^{-4}$ |

**S4 Table.** Exponential Fits to R. Same as S3 Table but for pandemic R(*t*_*p*_) values for the 49 U.S. jurisdictions.

| Parameter | Estimate | Standard Error | $Pr(> \vert t\vert )$ |
| --- | --- | --- | --- |
| $R_\infty$ | 0.97 | 0.024 | $< 2^{-16}$ |
| $R_0$ | 1.99 | 0.039 | $< 2^{-16}$ |
| $\alpha$ | 0.094 | 0.0017 | $< 2^{-16}$ |

**S5 Table.**
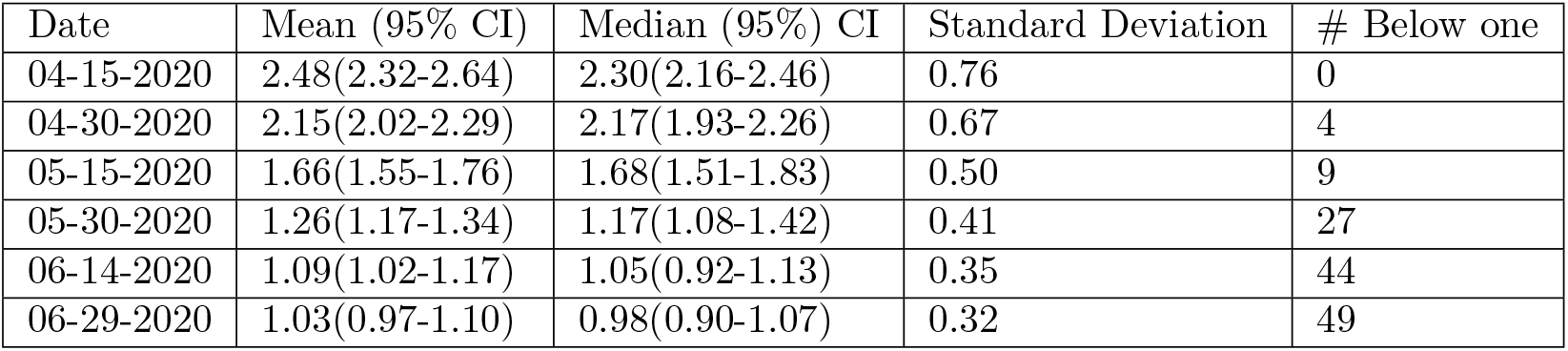
Calendar time reproduction number. Estimated R(*t*_*c*_) values for 89 global locations as a function of calendar time. Numbers in parentheses denote the 95% confidence interval of the mean and median values and SD denotes standard deviation.

| Date | Mean (95% CI) | Median (95% CI) | Standard Deviation | # Below one |
| --- | --- | --- | --- | --- |
| 04-15-2020 | 2.48(2.32-2.64) | 2.30(2.16-2.46) | 0.76 | 0 |
| 04-30-2020 | 2.15(2.02-2.29) | 2.17(1.93-2.26) | 0.67 | 4 |
| 05-15-2020 | 1.66(1.55-1.76) | 1.68(1.51-1.83) | 0.50 | 9 |
| 05-30-2020 | 1.26(1.17-1.34) | 1.17(1.08-1.42) | 0.41 | 27 |
| 06-14-2020 | 1.09(1.02-1.17) | 1.05(0.92-1.13) | 0.35 | 44 |
| 06-29-2020 | 1.03(0.97-1.10) | 0.98(0.90-1.07) | 0.32 | 49 |

**S6 Table.**
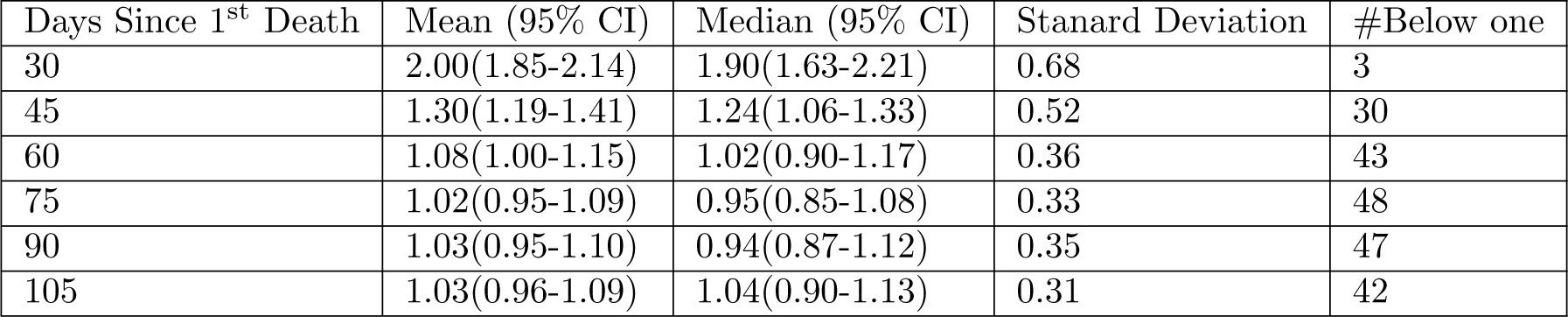
Pandemic time reproduction number. Same as S5 Table but as a function of local pandemic time, R(*t*_*p*_).

**S7 Table.** Exponential Fits to R. Results of exponential fit *R* (*t*) = *R*_∞_ + (*R*_0_ − *R*_∞_) *e*^−*αt*^ to calendar R(*t*_*c*_) values for the 89 global locations.

| Parameter | Estimate | Standard Error | $Pr(> \vert t\vert )$ |
| --- | --- | --- | --- |
| $R_\infty$ | 0.42 | 0.26 | 0.117 |
| $R_0$ | 2.55 | 0.052 | $< 2^{-16}$ |
| $\alpha$ | 0.018 | $4.4 * 10^{-4}$ | $< 2^{-16}$ |

**S8 Table.** Exponential Fits to R. Same as S7 Table but for pandemic R(*t*_*p*_) values for the 89 global locations.

| Parameter | Estimate | Standard Error | $Pr(> \vert t\vert )$ |
| --- | --- | --- | --- |
| $R_\infty$ | 1.01 | 0.030 | $< 2^{-16}$ |
| $R_0$ | 2.00 | 0.047 | $< 2^{-16}$ |
| $\alpha$ | 0.094 | 0.0014. | $< 2^{-16}$ |

## Notes

### Competing Interest Statement

The authors have declared no competing interest.

### Funding Statement

This work was supported by the 348 National Science Foundation Award number 2031536, and by Cooperative Agreement 349 number NU38OT000297 from The Centers for Disease Control and Prevention (CDC) 350
and CSTE and does not necessarily represent the views of CDC and CSTE.

